# Upper common pathways analysis using late atrial premature depolarization in atrioventricular nodal reentry tachycardia

**DOI:** 10.1101/2023.10.22.23296531

**Authors:** Soyoon Park, Jeong-Wook Park, Soohyun Kim, Hwajung Kim, Sung-Hwan Kim, Yong-Seog Oh, Young Choi

## Abstract

**Introduction:** The presence of an upper common pathway (UCP) in atrioventricular nodal reentrant tachycardia (AVNRT) has been suggested. However, the precise anatomical description and the actual prevalence of UCP have not been well established. We aimed to assess the prevalence of UCP in AVNRT using a late atrial premature depolarization (LAPD) maneuver.

**Methods:** Patients who were diagnosed with typical AVNRT by electrophysiologic studies were enrolled. To evaluate the presence of UCP, a LAPD was given at the coronary sinus ostium (osCS) during AVNRT and then pacing was repeated incrementally every 10 ms. The result was interpreted as i) absence of UCP, a LAPD from osCS can reset the tachycardia without depolarizing the earliest retrograde atrial activation site (ERAS) near the proximal His; ii) presence of UCP, a LAPD from osCS can depolarize the ERAS without resetting the tachycardia; and iii) indeterminate, a LAPD from osCS either reset the ERAS and tachycardia simultaneously or does not reset both.

**Results:** The LAPD maneuver was performed in 126 patients with AVNRT and one patient with nodoventricular orthodromic reentrant tachycardia (NVORT). The maneuver result demonstrated an absence of UCP in 121 (96.0%) patients, a presence of UCP in 3 (2.4%) patients, and was indeterminate in 2 (1.6%) patients. In the patient with NVORT, the result was consistent with the presence of UCP.

**Conclusions:** The LAPD maneuver revealed that most AVNRTs did not exhibit a single UCP. The presence of UCP was suggested in 2.4% of AVNRT cases and one case of NVORT.

## Introduction

It has been suggested that an AVNRT is an intranodal reentry composed of slow and fast pathways and the circuit is connected to atrial tissue by a single electrical connection, which is called the upper common pathway (UCP) (1–6). However, anatomical evidence demonstrates that the atrioventricular (AV) nodal tissue extends into the right and left atrium along the posterior septal tricuspid and mitral vestibules, forming a slow pathway (SP).(7,8) In addition, there is another connection in the anterior septum, which is the last septal input to the AV node. The last septal input is histologically working atrial cardiomyocytes, so it has a faster conduction velocity than SP and is therefore known as thefast pathway (FP) (9–12). The existence of multiple atrionodal connections is not compatible with the assertion that the circuit of AVNRT is solely connected to the atrium through a single UCP. Heidbuchel and Jackman have described an AVNRT circuit that includes an atrial myocardium component, as well as an AV node and ≥2 atrionodal connections (13).

Despite the absence of anatomical explanations, the belief in the existence of UCP is based on evidence that supports the electrical dissociation of AVNRT from the atrium; such as the characteristic of the ventriculo-atrial (VA) block during AVNRT, representatively (4). Meanwhile, Satoh et al. showed that burst atrial pacing during AVNRT could orthodromically capture atrial electrogram near the His bundle potential in 5 of 7 patients, which would not be anticipated if the atrium was not involved in the reentry circuit (14). This conflicting evidence suggests that the actual circuit of AVNRT may not be uniform in all cases. Miller et al. reported that UCPs were present in 8 of 28 (29%) patients with AVNRT, with a postulation that a discrepancy between atrial-His (AH) interval during AVNRT and atrial pacing suggests the presence of UCP (5). However, the prevalence of UCPs might have been overestimated, in that the maneuver could yield a false-positive result if the proximal AV junction catheter is not accurately positioned at the precise atrial exit site. Since then, most reports related to UCP have been small-scale studies providing evidence for either the presence or absence of UCPs (1–6).

The ability of late atrial premature depolarization (LAPD) delivered to the inferior atrial septum to advance the AVNRT implies the necessity of an atrium in the reentrant circuit and the absence of UCP (15). Meanwhile, depolarization of perinodal atrial tissue surrounding the AV node without affecting AVNRT indicates an intranodal location of reentry and the presence of UCP (4). With an observation of the reset response to a LAPD during AVNRT, the above two responses can be evaluated in the same instance. We sought to analyze the prevalence of UCPs in AVNRT using LAPDs delivered from the vicinity of slow pathway insertion, in a larger number of patients.

## Methods

We analyzed the data of patients with AVNRT who underwent electrophysiologic studies (EPS) between November 2019 and March 2023 in Seoul St. Mary’s Hospital, The Catholic University of Korea. The LAPD maneuver was performed in 126 patients who had sustained typical AVNRT during EPS. The LAPD maneuver was also performed in one other patient in whom typical AVNRT was suspected initially but nodoventricular orthodromic reentrant tachycardia (NVORT) was the final diagnosis (16). This study was approved by the Institutional Review Board (IRB) of Catholic Medical Center. The requirement for informed consent was waived by the IRB because we only used clinical and procedural data that was anonymously encoded.

### Electrophysiology studies

All antiarrhythmic drugs were discontinued at least 2 days before the procedure. Diagnostic mapping catheters were inserted through the femoral vein and positioned to the lateral side of the right atrium, coronary sinus, His area, and right ventricular apex. Electrograms were recorded using a digital recording system (CardioLab, GE Healthcare, Chicago, IL, USA). At baseline, ventricular and atrial programmed stimuli were delivered to identify the presence of accessory pathway and/or SP and then supraventricular tachycardia induction was attempted. If SVT was not induced, atrial and ventricular programmed stimuli were delivered again after intravenous isoproterenol infusion. Once supraventricular tachycardia was induced, His-refractory ventricular premature beat was delivered and overdrive pacing from the right ventricular apex was performed. The diagnosis of typical AVNRT was based on i) evidence of dual AV nodal physiology in the baseline study, ii) narrow QRS tachycardia with a VA interval < 90 ms, iii) absence of reset response to His-refractory ventricular premature beat, and iv) corrected post-pacing interval (PPI) – tachycardia cycle length > 115 ms after right ventricular overdrive pacing (17). After the diagnosis of AVNRT was confirmed, a LAPD maneuver was performed and slow pathway ablation was conducted. The ablation was targeted at the anatomical region of slow pathways and preferably, the sites showing sharp slow pathway electrograms. The ablation endpoint was a demonstration of spontaneous junctional rhythm, and the procedural success was defined as a non-inducibility of AVNRT or ≥2 echo beats.

### The LAPD maneuver

The LAPD maneuver assumes that if a single UCP is connecting the atrium and AVNRT circuit, an LAPD will not be able to invade the tachycardia circuit without capturing the atrial exit site of the UCP, which would be the earliest retrograde atrial activation site (ERAS) during AVNRT. The schematic description of the LAPD maneuver is presented in Figure 1. A LAPD was given at the coronary sinus ostium (osCS) during AVNRT and we assessed the reset response of the tachycardia and the ERAS. When the LAPD could not reset both tachycardia and ERAS, LAPD was given repeatedly in 10-ms increments. The result was interpreted as;

i. Absence of UCP: a LAPD from osCS can reset (or terminate) the tachycardia without depolarizing the ERAS near the proximal His (Figure 1A)
ii. Presence of UCP: a LAPD from osCS can depolarize the ERAS without resetting the tachycardia (Figure 1B)
iii. Indeterminate: a LAPD from osCS either resets the ERAS and tachycardia simultaneously or does not reset both.

**Figure 1.**
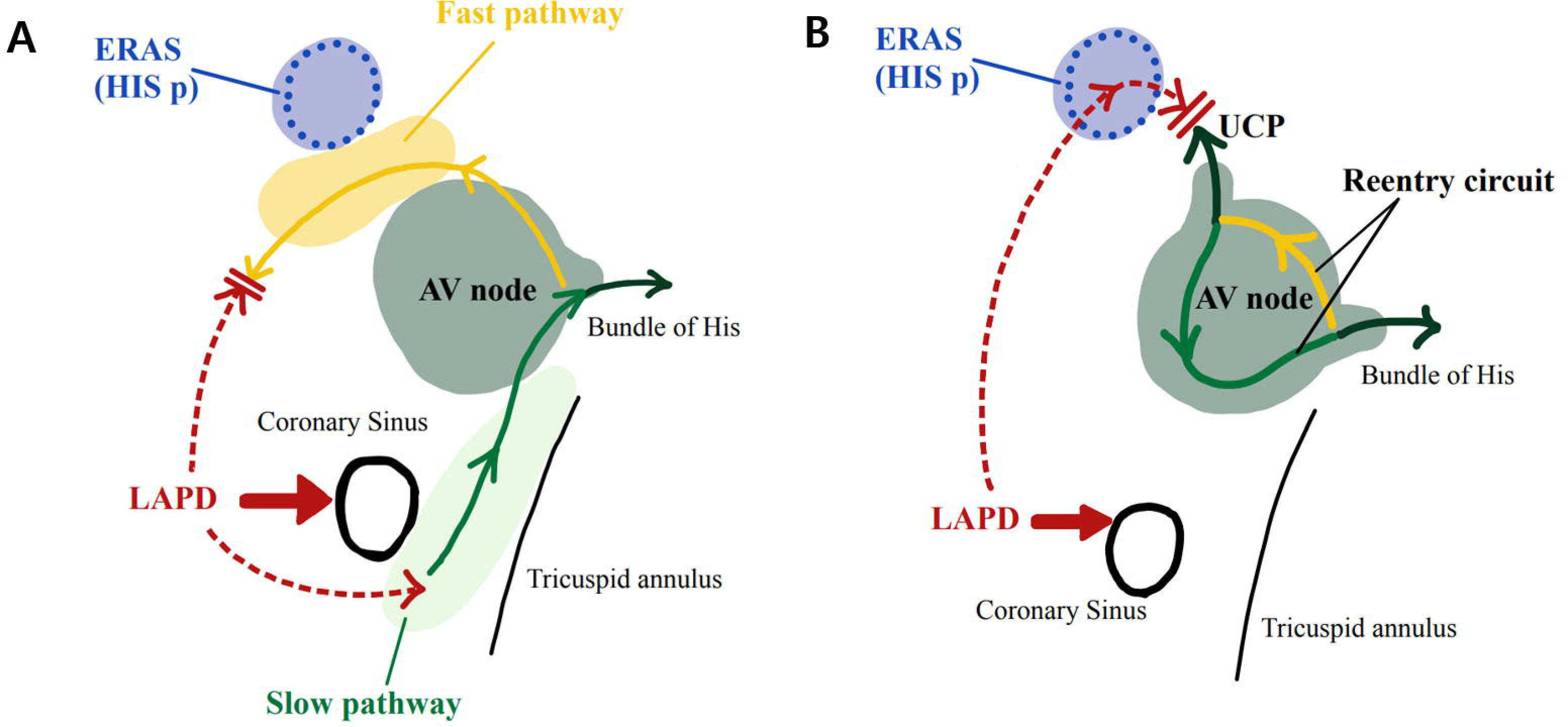
Schematic illustration of the LAPD maneuver. A) Absence of UCP response: If the AVNRT circuit involves the atrium, a LAPD from the coronary sinus ostium can enter the tachycardia circuit via the slow pathway without depolarizing the ERAS. B) Presence of UCP response: If there is a single connection between the atrium and intranodal reentrant circuit of AVNRT, the LAPD must first depolarize the ERAS. The LAPD would be able to depolarize the ERAS without resetting the tachycardia due to the gap generated by the UCP. LAPD = late atrial premature depolarization; UCP = upper common pathway; AVNRT = atrioventricular nodal reentrant tachycardia; ERAS = earliest retrograde atrial activation site

## Results

The LAPD maneuver was performed in 126 patients with typical AVNRT and one patient with NVORT. The mean age was 50.71±17.22 and 41 patients (32.8%) were male. Among the 126 patients who met the diagnostic criteria for typical AVNRT, an atypical AVNRT (fast/slow) was also induced in two patients, and the LAPD maneuver was performed during typical AVNRT. The LAPD maneuver results demonstrated an absence of UCP in 121 (96.0%) patients, a presence of UCP in 3 (2.4%) patients, and indeterminate in 2 (1.6%) patients (Figure 2). In two patients with indeterminate results, a repeated LAPD maneuver was performed but the same responses were obtained (ERAS and tachycardia were reset simultaneously). Among the 121 patients showing an absence of UCP response, the reset response was an advance of tachycardia in 118 patients and a termination of tachycardia in 3 patients. In a patient with NVORT, a short-RP tachycardia morphology was confused with typical AVNRT initially and the LAPD maneuver was performed. However, the His-refractory ventricular premature beat could delay the tachycardia, and the result from the ventricular entrainment showed very slow, decremental VA conduction and the first orthodromically captured His potential preceding the last entrained atrial electrogram, which confirms the diagnosis of NVORT (Figure 3A). In this patient, the LAPD maneuver result was consistent with the presence of UCP (Figure 3B).

**Figure 2.**
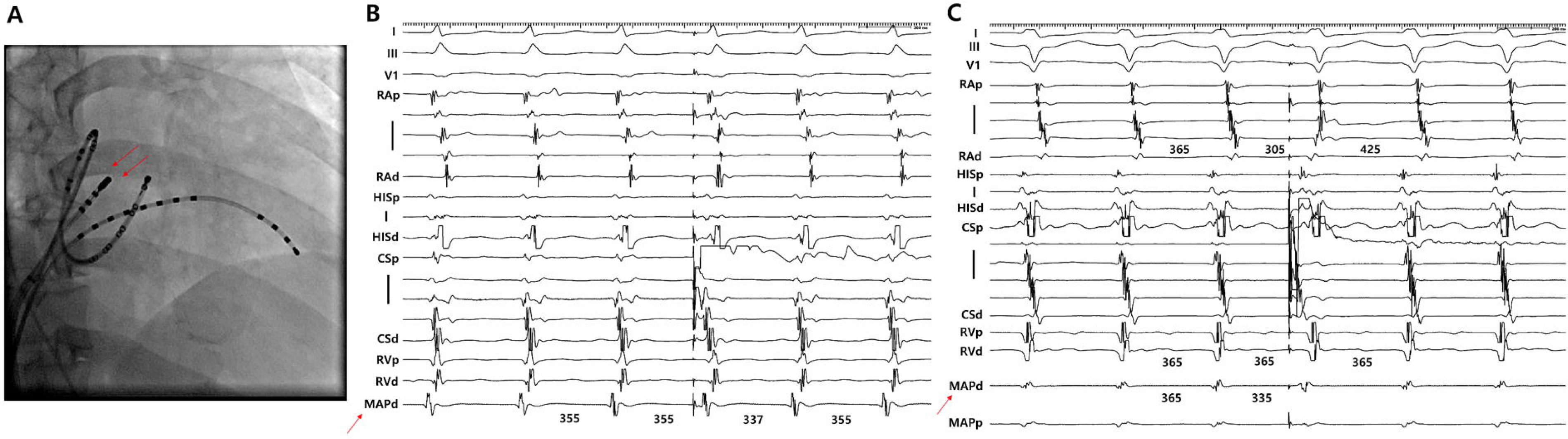
The LAPD maneuver results. A) Catheter position in RAO view. The mapping catheter is positioned at the ERAS (red arrows). B) Representative case of “absence of UCP” response. A LAPD delivered at the proximal CS during AVNRT did not immediately reset the ERAS (red arrow), but the next tachycardia beat was advanced. C) Representative case of “presence of UCP” response. A LAPD during AVNRT immediately reset entire atrial electrograms, including the ERAS (red arrow), CS, and RA electrograms without resetting the next tachycardia beat (RV electrograms). LAPD = late atrial premature depolarization; RAO = right anterior oblique; ERAS = earliest retrograde atrial activation site; UCP = upper common pathway; AVNRT = atrioventricular nodal reentrant tachycardia; CS = coronary sinus; RA = right atrium; RV = right ventricle

**Figure 3.**
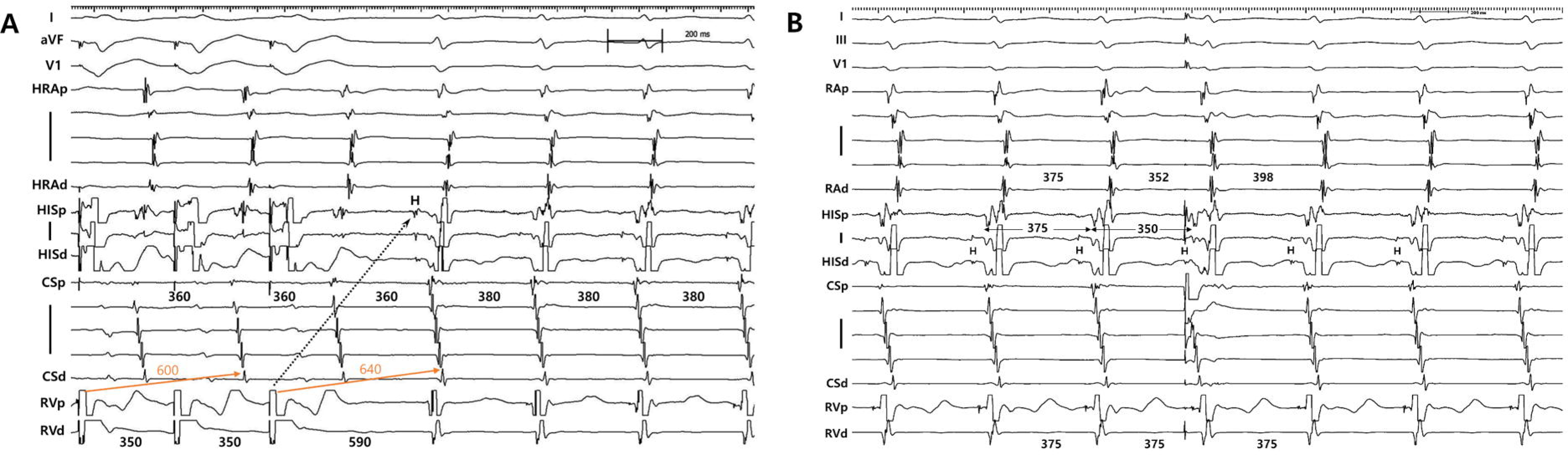
Maneuver results in a NVORT. A) Entrainment from the RV shows very slow decremental retrograde conduction (purple arrows) resulting in prolonged PPI. This is not expected in typical AVNRTs, and the first orthodromically captured His electrogram precedes the last entrained atrial electrogram, which excludes the possibility of AVNRT (dotted arrow). B) A LAPD delivered at the proximal CS during the NVORT resets the entire atrial electrograms including the ERAS near the His electrogram, but did not affect the next tachycardia beat, which is consistent with the “presence of UCP” response. NVORT = nodoventricular orthodromic reentrant tachycardia; RV = right ventricle; PPI = post-pacing interval; AVNRT = atrioventricular nodal reentrant tachycardia; CS = coronary sinus; ERAS = earliest retrograde atrial activation site; UCP = upper common pathway.

All patients underwent radiofrequency ablation for the right inferior slow pathway, and the ablation was successful, as defined by the achievement of non-inducible AVNRT or ≥2 echo beats. There was no intraprocedural complication. The successful ablation site was no different between patients with an absence or presence of UCP according to the LAPD maneuver. The patient with an NVORT also underwent radiofrequency ablation in the vicinity of the right inferior slow pathway area, and the tachycardia was non-inducible after the ablation.

## Discussion

In this study, we analyzed the prevalence of UCP in typical AVNRT using a simple maneuver that can exclude the presence of a single electrical connection between the atrium and the AV node during tachycardia. Most (96%) of the typical AVNRTs did not have a single UCP and the atrium (osCS area) was probably part of the reentry circuit. The presence of a single UCP was suggested in only 2.4% of typical AVNRTs. Additionally, we found that in an NVORT, the result of this maneuver was consistent with the presence of UCP.

The ability of LAPD delivered to the right intra-atrial septum to reset AVNRT was previously reported in a small case series (18). In this study, we recorded the ERAS electrogram to exclude the possibility of antidromic fast pathway activation and included a higher number of patients to assess whether this maneuver yields the same result in a large population. This maneuver has several advantages; it only requires a single premature beat with minimal concern about termination of tachycardia due to procedure. The observation of the reset responses in the ERAS and tachycardia by the premature beat given at a distant site from the AV node can yield two different results: resetting the tachycardia without affecting the ERAS excludes the possibility of intranodal reentry being connected to the atrium through UCP and resetting the ERAS without affecting the tachycardia strongly suggests the presence of UCP. With this maneuver, our study demonstrated that the actual prevalence of UCP is much lower than previously anticipated in the largest number of patients. This finding is consistent with a previous anatomical study about the spatial characterization of the slow or fast pathways, which is not consistent with a hypothesis that a single connection exists between the atrium and the AV node (12). Keim et *al*. localized the slow and fast pathways by intraoperative ICE mapping and demonstrated that atrial insertion sites of slow fibers are located apart from the AV node and were formed along the TV annulus (12). Moreover, cooling at the intermediate site between slow and fast pathways did not disturb sustaining the tachycardia, which provides evidence against a single connection composed of non-atrial tissue between fast and slow pathways during AVNRTs.

The hypothesis that a single UCP is present suggests that the atrium does not participate in the circuit of the AVNRT. This assumption has been supported by previous case reports, which indicated that AVNRT was able to persist despite the retrograde conduction block or VA dissociation during tachycardia (1,2,6). However, this phenomenon can be observed in only a very small subset of all AVNRT cases (19). In 1987, Miller et *al*. conducted a study to assess the prevalence of UCP (5). The study applied two criteria to define the presence of UCP: i) a difference between paced AH interval and the AH in SVT or ii) AV Wenchebach block during atrial pacing at the same cycle length as the tachycardia. With these criteria, they concluded that the prevalence of UCP was 29%. However, for the comparison of the AH interval during atrial pacing to that during SVT, there should be a premise that the atrial is recorded at the exact location where the atrium is connected to the AV node. If the AH interval was measured at a distant site from the AV node-atrium junction site, discrepancies can occur between the anterogradely and retrogradely activated AH intervals, resulting in a false-positive outcome. For the second criterion, the atrioventricular conduction during atrial pacing is expected to be the same as that during AVNRTs; but a wide variety of AV nodal conduction delay or block response can occur depending on the pattern of atrial input, and intrinsic or extrinsic influences (20). In our study, the presence of a UCP was not suggested in most patients, but 3 cases (2.4%) appeared to have UCPs and the maneuver results were difficult to interpret in 2 cases. In all 3 cases where a UCP was indicated, nearly all atrial electrograms, including the ERAS, were reset by the LAPD maneuver, whereas the tachycardia was not. This criterion is known to suggest the presence of UCP (4). Nevertheless, even in the absence of a UCP, if the pacing site is distant from the insertion of the slow pathway, there is a possibility that this maneuver could yield a false-negative result. Additionally, a delay in the slow pathway conduction could have been compensated by the prematurity of atrial depolarization, resulting in the tachycardia cycle length becoming balanced with the previous beat (21). Therefore, it would be cautious to interpret the results of our study findings as definitively indicating the presence of a UCP in a certain proportion of AVNRTs.

Recent studies have described the characteristics of concealed nodoventricular bypass tract (NVBT) or nodofascicular bypass tract (NFBT), which were not well-recognized previously (22–24). The presence of concealed NVBT/NFBT as bystanders in AVNRT, or orthodromic reciprocating tachycardias (ORT) using these bypass tracts can confuse the diagnosis and lead to the incorrect interpretation of a VA block as if there is a UCP in AVNRT (25). However, the ORTs using NVBT or NFBT are difficult to recognize by simple electrophysiologic study maneuvers (26). To discriminate the tachycardias from AVNRT, a response to His-refractory PVC, PPI from the RV, differential pacing at the RV, and the analyses of QRS complex fusion during entrainment could be performed (26). The most reliable maneuver is PPI; however, the optimal cutoff value to discriminate AVNRTs from nodoventricular (NV)/nodofascicular (NF) ORTs is not clear, and PPI can be unexpectedly prolonged in NV/NFORTs due to a decremental property in the bypass tracts (27). Therefore, even with the implementation of standard electrophysiological maneuvers, there remains a chance that VA-dissociated NV/NFORTs might be misidentified as an AVNRT with a UCP. Comprehensive studies for precise differentiation are imperative when a narrow QRS tachycardia with VA dissociation is demonstrated; nonetheless, the actual effective ablation sites for both tachycardias are similar, and it is possible that a significant number of cases are not properly diagnosed but are still successfully treated (24). The LAPD maneuver used in our study suggests the atrium is a part of the AVNRT circuit in most cases; additionally, the maneuver could differentiate an NVORT from AVNRTs, in which the atrium has no role in maintaining tachycardia. In NV/NFORTs, there could be dual connections between the atrium and AV node; however, both fast and slow pathways are activated retrogradely during tachycardia, unlike AVNRTs. In such situations, the LAPD would not be able to penetrate the AV node primarily via the slow pathway and reset the tachycardia without activating the fast pathway. The diagnostic value of this maneuver in distinguishing NV/NFORTs from AVNRTs should be validated further in a larger number of cases.

### Limitations

The LAPD maneuver is not effective in confirming the presence of a UCP. A positive result is defined by the ability of the LAPD to penetrate the tachycardia without depolarizing the ERAS. However, if the pacing impulse is delivered far from the slow pathway insertion, it may not be able to penetrate the tachycardia before reaching the ERAS, even in the absence of a UCP. To mitigate this potential error, we limited the analysis to patients who were confirmed to have slow pathways of the right inferior extension. Nevertheless, the limitation of the maneuver regarding false negatives could not be fully addressed. Second, this maneuver could not be applied in atypical AVNRTs, because the ERAS was only consistent in tachycardias using a fast pathway as a retrograde limb. In this maneuver, it is crucial to accurately record the electrogram of the ERAS, so the maneuver was only conducted during short-RP tachycardias. Third, as discussed above, distinguishing between NV/NFORT and AVNRT can be challenging, and the response to the LAPD maneuver varies between these different tachycardias. In this study, AVNRT was diagnosed using PPI; however, the accuracy of PPI in the exclusion of NV/NFORT is not yet clear (27). For this reason, we performed a detailed review of the electrogram in 3 cases showing positive UCP responses among AVNRTs. The results from the response to His-refractory PVC and differential RV entrainment suggest that they were not compatible with NV/NFORTs.

## Conclusion

We analyzed the prevalence of UCPs in 126 patients with typical AVNRTs. A single UCP was absent in 96% of patients, and the presence of UCP was suggested in only 2.4%. In addition, when we applied this maneuver to a patient with an NVORT, which is definitely an intranodal reentrant tachycardia, the response was consistent with the presence of a UCP. Although previous cases displaying VA dissociation during AVNRT have implied the presence of UCP and an intranodal reentrant circuit, our findings indicate that the atrium is involved in the reentry circuit of most AVNRTs. The potential efficacy of the LAPD maneuver in distinguishing AVNRTs from other intra or infranodal tachycardias should be validated in future research.

## Abbreviations

AVNRT: Atrioventricular nodal reentry tachycardia
LAPD: Late atrial premature depolarization
osCS: Coronary sinus ostium
UCP: Upper common pathway
ERAS: Earliest retrograde atrial activation site
SP: Slow pathway
FP: Fast pathway
ORT: Orthodromic reciprocating tachycardias
NVORT: Nodoventricular orthodromic reentrant tachycardia
NFORT: Nodofascicular orthodromic reentrant tachycardia

## Data Availability

All data produced in the present study are available upon reasonable request to the authors

## Notes

### Competing Interest Statement

The authors have declared no competing interest.

### Funding Statement

This study did not receive any funding

### Author Declarations

This study was approved by the Institutional Review Board (IRB) of Catholic Medical Center. The requirement for informed consent was waived by the IRB because we only used clinical and procedural data that was anonymously encoded.

